# From Conversation to Chart: An Analysis of Clinician Edits to Ambient AI Draft Notes

**DOI:** 10.64898/2026.01.05.26343471

**Authors:** Yawen Guo, Di Hu, Yiliang Zhou, Tianchu Lyu, Sairam Sutari, Steven Tam, Emilie Chow, Danielle Perret, Deepti Pandita, Kai Zheng

## Abstract

**Objective:** Ambient artificial intelligence (AI) tools are increasingly adopted in clinical practices. This study investigated whether and how clinicians edit AI-generated drafts and the linguistic differences between AI drafts and clinician-finalized notes.

**Materials and Methods:** This retrospective study analyzed real-world data from ambulatory clinics at a large academic health system spanning two vendor deployments. We quantified clinicians’ editing behavior using the Myers diff algorithm to compare AI drafts and final documentation. We then applied statistical and linguistic analysis to study factors associated with the frequency/intensity of editing across note sections, turnaround time, clinician characteristics, and encounter types.

**Results:** Across 23,760 notes that included one or more ambient AI sections, 84.4% were edited by clinicians before signing off. While rates of unedited notes differed across note sections and care settings, the dominant source of variation was individual clinician practice style rather than specialty-level norms. Notes signed after 24 hours had lower overall edit intensity. The final versions showed small but statistically significant linguistic changes and exhibited slightly higher lexical diversity and modest changes in readability. Editing is most intensive in the assessment and plan section, and varies across specialties.

**Conclusion and Discussion:** A majority of AI-drafted clinical notes were edited by clinicians, although the editing rate varies across note sections, medical specialties, and individual clinicians. Future research is needed to further analyze this editing behavior to inform improvement in AI-assisted clinical documentation to achieve better documentation quality, efficiency, and clinician satisfaction.

## Background and Significance

Clinical documentation is essential to outpatient care but remains a major driver of after-hours “pajama time” and clinician burnout. [1–3] Ambient artificial intelligence (AI) documentation systems have emerged to capture visit conversations and draft notes that integrate into the electronic health record (EHR) for clinicians to review. [4,5] Prior studies report that clinicians are generally satisfied with ambient AI tools, citing reduced documentation burden and improvements in workload, work–life integration, and patient engagement. [6–10] Despite this promise, clinicians still spend substantial time reviewing, refining, and signing off notes, and analysis on time savings alone do not reveal how clinicians adopt and modify AI drafts in routine practice. Most evaluations of ambient AI documentation to date rely on before-and-after implementation comparisons, assessing time-based workflow efficiency outcomes such as time spent on note writing. [11–13] Early evidence suggests that following deployment, clinicians spend less time on documentation while AI-assisted notes become longer, raising concerns about note bloat, in which increased length may dilute clinically important content and increase downstream review burden. [6,14]

In contrast, routine editing behavior in real-world practice remains poorly characterized. It is still unclear whether clinicians sign ambient AI-drafted notes without changes, how they revise drafts, and how editing behaviors vary by different encounter characteristics. Empirical evidence is also limited in how ambient AI relates to note completion timeliness within everyday ambulatory workflows. Understanding these patterns is essential because documentation practices and information needs differ substantially across clinical settings such as specialties and visit types, and ambient AI documentation should support workflow-specific needs without shifting effort into additional review and correction work. [2,15,16]

To address this gap, we analyzed a large dataset of paired ambient AI–drafted note and clinician-finalized text from a large academic health system, where two commercially available ambient AI documentation systems are integrated into ambulatory EHR workflows. Our analyses addressed three questions: (1) how often AI drafts are submitted without edits and whether no-edit behavior reflects individual clinician writing style versus specialty norms, as well as how it varies by note section, encounter type, and specialty; (2) among edited note sections, how clinician-edited and AI-generated texts differ in linguistic characteristics, including length, lexical diversity, and readability, and how the magnitude of edits varies by note sections and care settings; and (3) how turnaround time relates to edit intensity, and which note sections, encounter types, and specialties are associated with more editing and longer turnaround. Together, these analyses provide a detailed picture of how clinicians interact with ambient AI documentation in routine outpatient practice.

## Methods

### Setting, Data Source, and Unit of Analysis

A pilot deployment of ambient AI documentation was initiated at University of California, Irvine Medical Center (UCI Health) in 2023. UCI Health is an academic health system with ambulatory clinics across multiple sites in Southern California. Participating physicians from selected primary care and specialty clinics used two commercially available ambient AI tools integrated with the Epic EHR. Vendors were referred as Vendor A and Vendor B to align with institutional data governance and reporting constraints. Clinicians accessed the tools through Haiku mobile application or through a browser-based interface connected to Epic. [17] In all workflows, AI-generated text was reviewed, edited as needed, and signed within the EHR, and clinicians retained full responsibility for the final documentation. [18] Clinicians received onboarding and workflow guidance as part of the pilot rollout, with ongoing technical support available during the study period. Consistent with institutional procedures for the pilot, clinicians informed patients at the visit that audio-assisted note drafting might be used, that the resulting documentation would be clinician-reviewed before filing, and that identifiable audio was not retained. Liability and regulatory considerations were managed through institutional compliance and IT review, and the EHR audit trail preserved both the initial AI-generated draft and the final signed version. We report overall pooled summaries and vendor-stratified analyses where applicable and do not disclose the mapping between the vendor labels.

We extracted all ambient AI note sections from Epic between September 2024 and August 2025. In routine clinical use, ambient AI generates draft text at the note-section level, and documentation is completed and signed at the encounter note level. Clinicians can selectively choose one or more AI-drafted note sections to insert into an EHR note. Four types of note sections can be generated by the ambient AI tools, including History of Present Illness (HPI), Assessment & Plan (A&P), Physical Exam, and Results. We analyzed workflow outcomes that are naturally defined per encounter (e.g., turnaround time and encounter-level unedited use) at the note level by aggregating section-level data within each unique note ID, and then conducted section-level analyses to characterize where edits concentrate across note-section types.

### Prevalence of Unedited Note Sections by Clinical Context

To quantify how often ambient AI drafts were accepted as-is, we first computed a note-level no-edit indicator (1 = all AI-generated sections within a note were signed without edits; 0 = otherwise) and summarized the prevalence of unedited use. We then conducted a section-level drill-down by defining a binary section-level indicator (1 = no-edit, 0 = edited) and stratifying unedited prevalence by note section, care setting (primary vs specialty care), and encounter type. For each dimension, we used χ² tests of independence to compare the distribution of unedited vs edited sections across categories, reporting χ² statistics, degrees of freedom, and p-values.[19] We also conducted vendor-stratified χ² tests as sensitivity analyses.

### Unedited Use as Personal Style vs Specialty Norms

To distinguish individual documentation style from broader specialty norms, we fit mixed-effects logistic regression models separately within each vendor, restricted to clinicians who have used ambient AI to draft over 100 note sections.[20] The binary outcome indicated whether a section was submitted without edits. Fixed effects included note section, encounter type, and clinician group (specialty vs primary care). We specified random intercepts for individual clinician and specific medical specialty (e.g., cardiology, neurology, etc.). [21] Including clinician group as a fixed effect captures broad differences between primary and specialty care, while the specialty random intercept captures residual variation across specific specialties beyond that grouping. We summarized variance attributable to each level by computing intraclass correlation coefficients (ICCs).

### Linguistic Metrics Before and After Editing

For note sections that were edited, we computed eight metrics for both AI draft and final version: character count, word count, sentence count, unique word count, type–token ratio (unique words divided by total words), average word length, Flesch Reading Ease, and Flesch–Kincaid Grade Level. [22] Within each note section we calculated a before–after difference for each metric by subtracting the AI draft from the clinician-final text (Δ = Final − AI). To quantify overall editing, we used paired t-tests to test whether the mean Δ differed from zero. [23] Within each vendor, we used paired t-tests to assess whether mean Δ differed from zero. To compare editing magnitude between vendors, we compared mean Δ between Vendor A and Vendor B using Welch’s two-sample t-test. [24] To examine variation by note section, we compared Δ across HPI, A&P, Physical Exam, and Results using one-way ANOVA with Tukey’s HSD post-hoc tests for pairwise contrasts. [25]

### Quantifying Edit Intensity with Myers Diff

For all edited sections, we aligned the AI draft and final note using a token-level Myers diff algorithm. [26] This approach measures edits at the token level, where each token corresponds to a word or punctuation unit. We defined total edit intensity as the total number of tokens changed, calculated as the sum of inserted, deleted, and replaced tokens.

### Factors Associated With Longer Turnaround Time

To identify which notes were more likely to have turnaround over 24 hours, we fit a logistic regression model using the binary turnaround indicator as the outcome, and encounter type, clinician specialty, and the transformed number of AI-generated note sections included in each note as predictors. Vendor type was included as a covariate, and we tested whether the association between section count and long turnaround differed by vendor using the vendor-by-log(1+sections) interaction by comparing nested models using a likelihood-ratio test. Encounter types and specialties with fewer than 20 notes, or with 0% or 100% turnaround prevalence over 24 hours, were collapsed into pooled “Other” categories. [27]

### Turnaround Time and Editing Intensity

A binary turnaround indicator was coded as 1 when time from AI draft generation to note signing exceeded 24 hours, and 0 otherwise. Because signing occurs at the note level and the draft and signing dates were identical across sections within a note, we conducted turnaround analyses at the note level by aggregating section-level edits within each note. To test whether later submissions received more edits, we first compared total note-level edit intensity (sum of inserted, deleted, and replaced tokens across all sections in the note) between notes signed within 24 hours and those signed after 24 hours using descriptive statistics and a Mann–Whitney U test. [28] We repeated these comparisons stratified by vendors. For clinicians who have used ambient AI to draft over 100 note sections, we then fit a mixed-effects model with log-transformed total edit intensity per note as the outcome and turnaround time indicator as the primary predictor, adjusting for encounter type, clinician group (primary vs specialty care), and the log-transformed number of AI-generated note sections included in each note as fixed effects, and including an individual clinician-level random intercept. [29] To avoid unstable estimates due to sparse categories or perfect prediction, encounter types, specialties, and section categories with small counts were collapsed into an “Other” category prior to modeling.

### Factors Associated With Heavier Editing Intensity

To examine which note sections, encounter types, and specialties were associated with heavier editing, we fit a regression model with log-transformed edit intensity as the outcome, and note section, encounter type, and clinician specialty as categorical predictors, including vendor type as a covariate, and using clinician-clustered robust standard errors to account for within-clinician correlation (i.e., clustering on clinician to allow correlation among sections authored by the same clinician). [30] Encounter types and specialties with fewer than 50 sections were pooled into “Other” categories.

### Ethical Oversight and Data Security

The work was approved by the University of California, Irvine Institutional Review Board (IRB #7123). All data processing occurred within HIPAA-compliant secure environments, including UCI Health’s Protected Virtual Computing Environment (PVCE) and Amazon Web Services (AWS) HIPAA-aligned infrastructure.

## Results

### Dataset Characteristics

The dataset included 23,760 unique notes containing 48,155 AI-drafted note sections, authored by 225 clinicians for 16,520 patients (Table 1). We summarized characteristics at the note-section level because clinicians can selectively insert and edit individual AI-generated smart elements, and a single note may contain multiple AI-drafted sections. When stratified by vendor, Vendor A contributed 12,439 notes and Vendor B contributed 11,321 notes. Full vendor-stratified cohort characteristics are provided in Table 1.

**Table 1.**
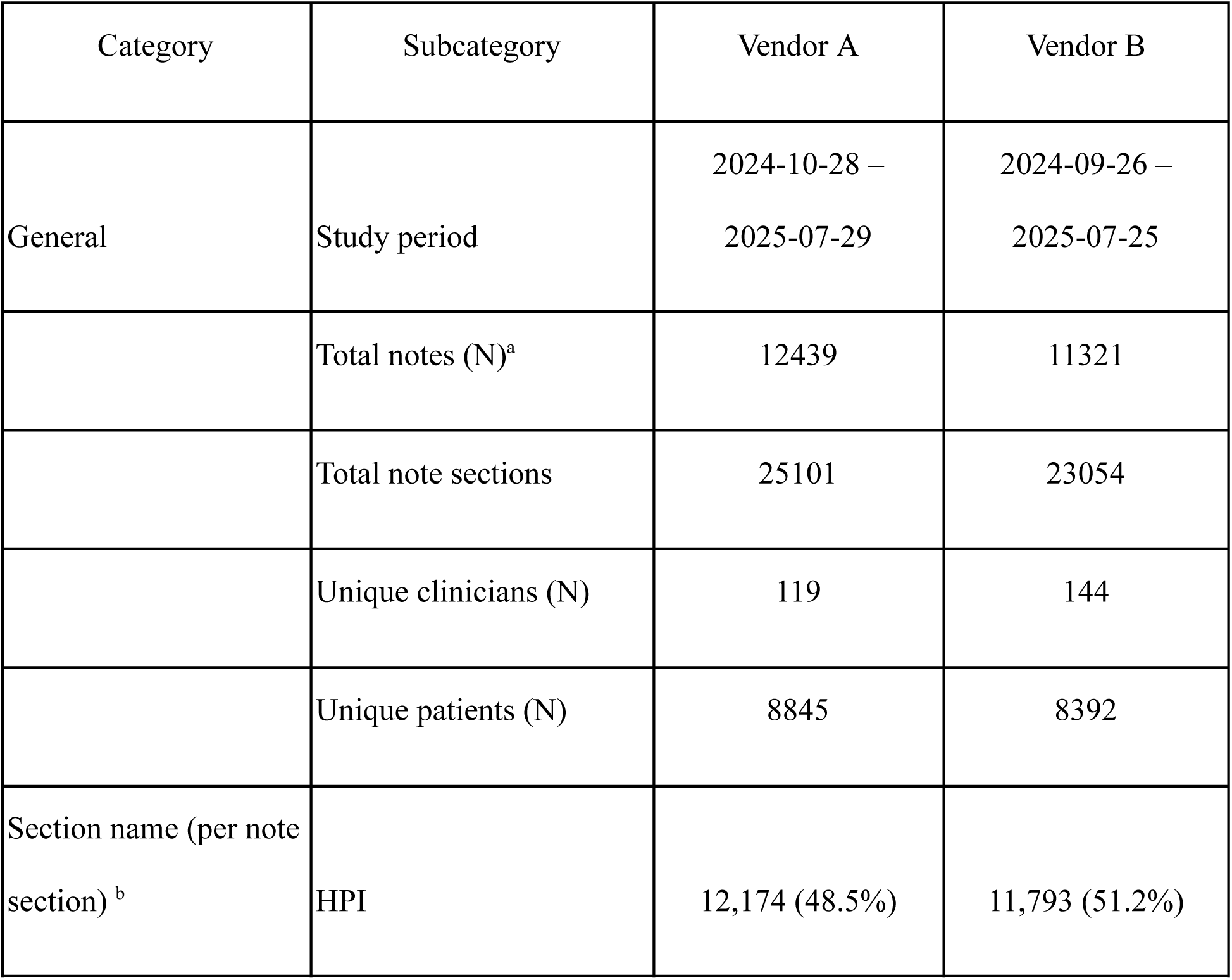
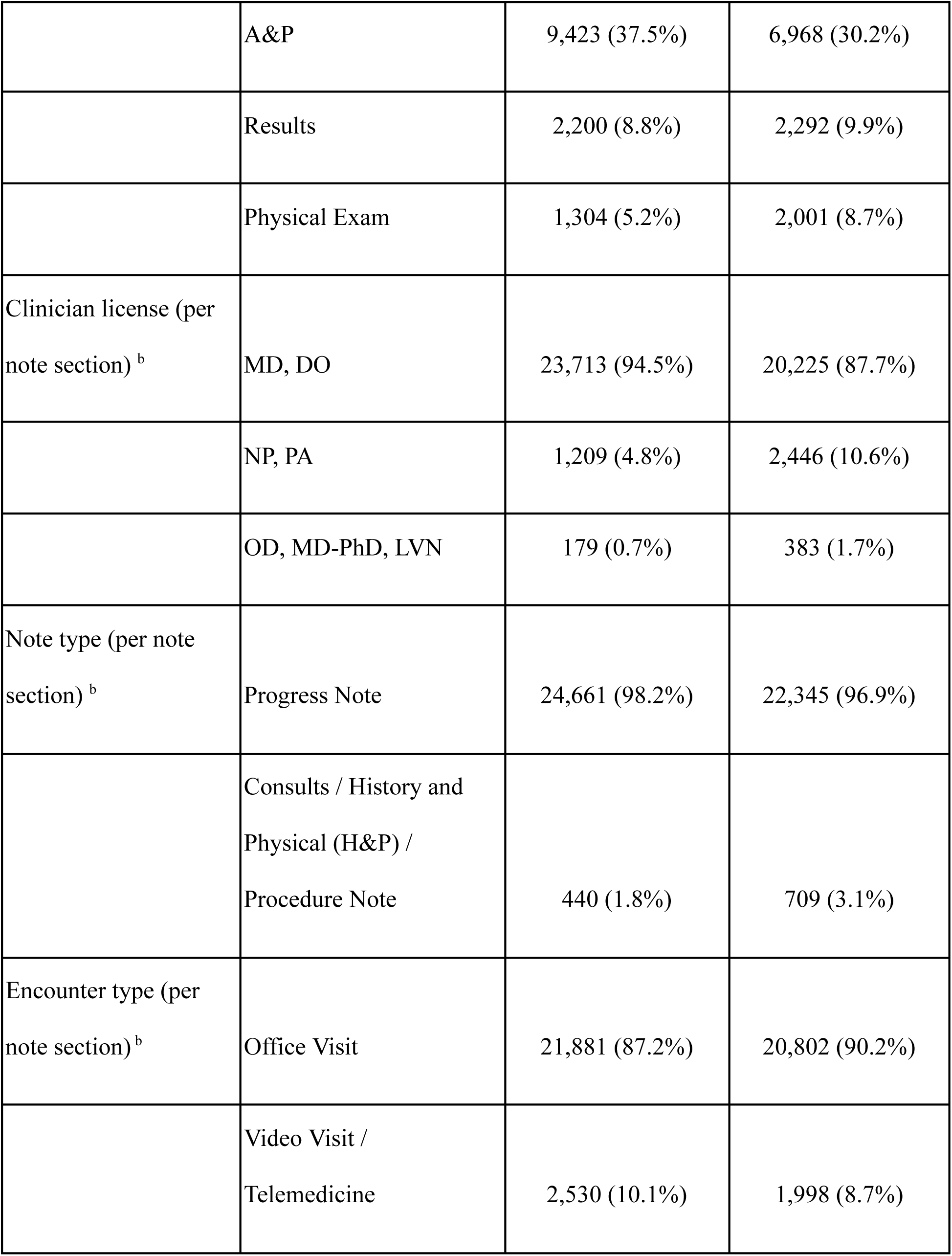
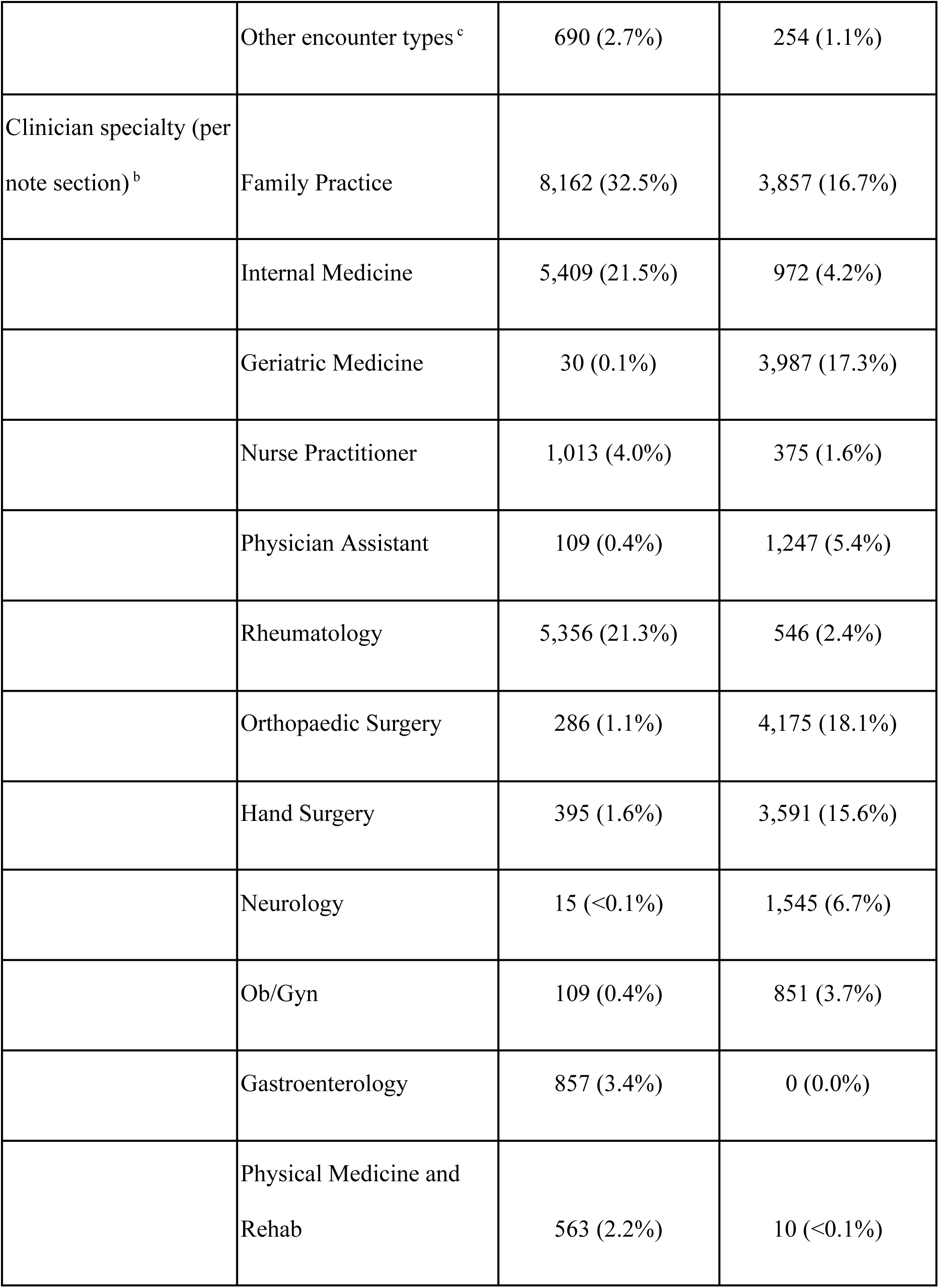
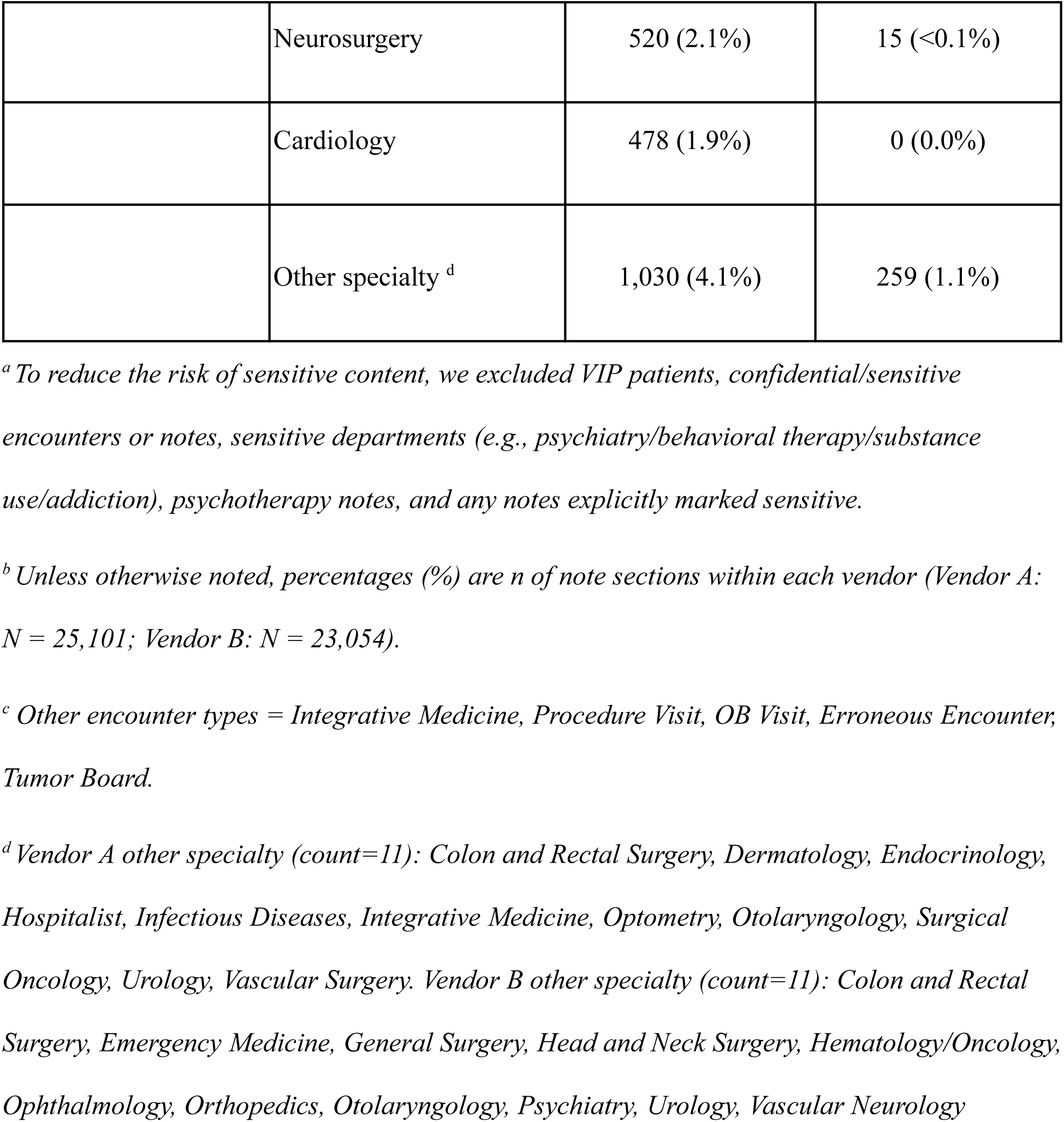
Study Cohort and Dataset Characteristics.

| Category | Subcategory | Vendor A | Vendor B |
| --- | --- | --- | --- |
| General | Study period | 2024-10-28 –<br>2025-07-29 | 2024-09-26 –<br>2025-07-25 |
|  | Total notes (N)<sup>a</sup> | 12439 | 11321 |
|  | Total note sections | 25101 | 23054 |
|  | Unique clinicians (N) | 119 | 144 |
|  | Unique patients (N) | 8845 | 8392 |
| Section name (per note section)<sup>b</sup> | HPI | 12,174 (48.5%) | 11,793 (51.2%) |
|  | A&P | 9,423 (37.5%) | 6,968 (30.2%) |
|  | Results | 2,200 (8.8%) | 2,292 (9.9%) |
|  | Physical Exam | 1,304 (5.2%) | 2,001 (8.7%) |
| Clinician license (per<br>note section) <sup>b</sup> | MD, DO | 23,713 (94.5%) | 20,225 (87.7%) |
|  | NP, PA | 1,209 (4.8%) | 2,446 (10.6%) |
|  | OD, MD-PhD, LVN | 179 (0.7%) | 383 (1.7%) |
| Note type (per note<br>section) <sup>b</sup> | Progress Note | 24,661 (98.2%) | 22,345 (96.9%) |
|  | Consults / History and<br>Physical (H&P) /<br>Procedure Note | 440 (1.8%) | 709 (3.1%) |
| Encounter type (per<br>note section) <sup>b</sup> | Office Visit | 21,881 (87.2%) | 20,802 (90.2%) |
|  | Video Visit /<br>Telemedicine | 2,530 (10.1%) | 1,998 (8.7%) |
|  | Other encounter types <sup>c</sup> | 690 (2.7%) | 254 (1.1%) |
| Clinician specialty (per<br>note section) <sup>b</sup> | Family Practice | 8,162 (32.5%) | 3,857 (16.7%) |
|  | Internal Medicine | 5,409 (21.5%) | 972 (4.2%) |
|  | Geriatric Medicine | 30 (0.1%) | 3,987 (17.3%) |
|  | Nurse Practitioner | 1,013 (4.0%) | 375 (1.6%) |
|  | Physician Assistant | 109 (0.4%) | 1,247 (5.4%) |
|  | Rheumatology | 5,356 (21.3%) | 546 (2.4%) |
|  | Orthopaedic Surgery | 286 (1.1%) | 4,175 (18.1%) |
|  | Hand Surgery | 395 (1.6%) | 3,591 (15.6%) |
|  | Neurology | 15 (<0.1%) | 1,545 (6.7%) |
|  | Ob/Gyn | 109 (0.4%) | 851 (3.7%) |
|  | Gastroenterology | 857 (3.4%) | 0 (0.0%) |
|  | Physical Medicine and<br>Rehab | 563 (2.2%) | 10 (<0.1%) |
|  | Neurosurgery | 520 (2.1%) | 15 (<0.1%) |
|  | Cardiology | 478 (1.9%) | 0 (0.0%) |
|  | Other specialty <sup>d</sup> | 1,030 (4.1%) | 259 (1.1%) |

<sup>a</sup> To reduce the risk of sensitive content, we excluded VIP patients, confidential/sensitive encounters or notes, sensitive departments (e.g., psychiatry/behavioral therapy/substance use/addiction), psychotherapy notes, and any notes explicitly marked sensitive.
<sup>b</sup> Unless otherwise noted, percentages (%) are n of note sections within each vendor (Vendor A: N = 25,101; Vendor B: N = 23,054).
<sup>c</sup> Other encounter types = Integrative Medicine, Procedure Visit, OB Visit, Erroneous Encounter, Tumor Board.
<sup>d</sup> Vendor A other specialty (count=11): Colon and Rectal Surgery, Dermatology, Endocrinology, Hospitalist, Infectious Diseases, Integrative Medicine, Optometry, Otolaryngology, Surgical Oncology, Urology, Vascular Surgery. Vendor B other specialty (count=11): Colon and Rectal Surgery, Emergency Medicine, General Surgery, Head and Neck Surgery, Hematology/Oncology, Ophthalmology, Orthopedics, Otolaryngology, Psychiatry, Urology, Vascular Neurology

### How Often Are AI Drafts Signed Without Edits?

Overall, 15.6% of the notes were signed without any edits (3,698/23,760), with slightly higher no-edit rates for Vendor A than Vendor B (17.4%, 2,164/12,439 vs 13.6%, 1,534/11,321). Across all note sections, 14,568 of 48,155 sections (30.3%) were signed without any provider-edited text; section-level no-edit rates were likewise higher for Vendor A than Vendor B (32.0%, 8,020/25,101 vs 28.4%, 6,548/23,054). Within each vendor, no-edit rate differed substantially by note section (Vendor A: χ²(3)=4,686.2, p<0.001; Vendor B: χ²(3)=1,794.8, p<0.001). For Vendor A, no-edit rates were 8.7% for A&P to 40.2% for HPI, 54.3% for Physical Exam, and 72.5% for Results; for Vendor B, rates were 19.2% for A&P to 25.1% for HPI, 41.6% for Physical Exam, and 62.0% for Results (Table 2). No-edit rates also varied by encounter type within both vendors (Vendor A: χ²(7)=152.9, p<0.001; Vendor B: χ²(5)=55.2, p<0.001). Office visits had a 33.0% no-edit rate for Vendor A and 28.4% for Vendor B; video visits were lower (26.3% and 24.2%), while telemedicine encounters were 32.2% and 36.0%, respectively. Differences by clinician group were significant for Vendor A (primary care 36.2% vs specialty care 26.1%; χ²(1)=275.5, p<0.001) but not for Vendor B (29.6% vs 29.1%; χ²(1)=0.7, p=0.418). We conducted a supplementary specialty-level analysis and found substantial heterogeneity in section-level no-edit rates across clinician specialties overall (χ²(33)=3528.4, p<0.001) and within each vendor when analyzed separately (Supplementary Table S1). Vendor-stratified supplementary analyses use the analytic subset with Vendor A/B mapping and complete covariates; denominators therefore differ from full cohort totals in Table 1.

**Table 2.** Section-Level No-Edit Rates by Clinician Group, Encounter Type, and Note Section.

| Dimension | Category | Vendor A |  | Vendor B |  |
| --- | --- | --- | --- | --- | --- |
|  |  | Total sections, n | No-edit sections, n (% within Vendor ) | Total sections, n | No-edit sections, n (% within Vendor) |
| Clinician<br>group | Primary Care | 14,723 | 5,323 (36.2%) | 10,438 | 3,093 (29.6%) |
|  | Specialty<br>Care | 10,046 | 2,624 (26.1%) | 11,239 | 3,273 (29.1%) |
| Encounter<br>type | Office Visit | 21,881 | 7,221 (33.0%) | 20,802 | 5,917 (28.4%) |
|  | Video Visit | 1,824 | 479 (26.3%) | 1,118 | 270 (24.2%) |
|  | Telemedicine | 706 | 227 (32.2%) | 880 | 317 (36.0%) |
|  | Integrative<br>Medicine | 614 | 77 (12.5%) | 0 | 0 (0.0%) |
|  | Procedure<br>Visit | 58 | 12 (20.7%) | 155 | 27 (17.4%) |
|  | OB Visit | 9 | 2 (22.2%) | 94 | 14 (14.9%) |
|  | Erroneous<br>Encounter | 3 | 2 (66.7%) | 5 | 3 (60.0%) |
|  | Tumor Board | 6 | 0 (0.0%) | 0 | 0 (0.0%) |
| Note section | HPI | 12,174 | 4,894 (40.2%) | 11,793 | 2,960 (25.1%) |
|  | A&P | 9,423 | 822 (8.7%) | 6,968 | 1,336 (19.2%) |
|  | Results | 2,200 | 1,596 (72.5%) | 2,292 | 1,420 (62.0%) |
|  | Physical Exam | 1,304 | 708 (54.3%) | 2,001 | 832 (41.6%) |

*\* Categories with 0 total sections in a vendor stratum were excluded from $\chi^2$ tests*
*\* Percentages are calculated within each vendor and category: no-edit sections / total sections for that vendor.*

### Is “no-edit” behavior driven by individual writing styles or specialty norms?

Unedited submissions were driven more by individual clinician style than by specialty norms in both vendor analysis. In mixed-effects logistic regression models fit separately for Vendor A and Vendor B (adjusting for note section and encounter type), specialty-care clinicians had slightly lower odds of signing AI note drafts without edits than primary-care clinicians (Vendor A: β_specialty = −0.13; Vendor B: β_specialty = −0.14). Random-effects variance was concentrated at the clinician level in both vendors (Vendor A: Var_clinician = 3.84, ICC = 0.54; Vendor B: Var_clinician = 2.47, ICC = 0.43), with negligible variance attributable to specialty (Vendor A: Var_specialty = 0.0023, ICC ≈ 0.00; Vendor B: Var_specialty = 0.0018, ICC ≈ 0.00). These findings indicate that “no-edit” behavior varied primarily across individual clinicians rather than across specialties after accounting for section type and encounter context (Supplementary Table S2).

### Linguistic Differences Between AI Drafts and Final Note Sections

Paired comparisons showed small but statistically significant differences between AI drafts and final note sections across most linguistic metrics (Table 3). In the vendor-stratified analysis, Table 3 summarizes 33,502 paired AI–final section comparisons in total (Vendor A: 16,996; Vendor B: 16,506). Across both vendors, final notes generally had fewer sentences and changes in readability and lexical diversity, although the magnitude and direction of changes varied by vendor. We additionally tested whether the before–after change (Final–AI) differed between vendors using a Welch two-sample t-test (i.e., a difference-in-differences comparison of mean deltas): the vendor gap in mean deltas was significant for most metrics (p<0.001), but not for sentence count (p=0.455) and only marginal for unique word count (p=0.071).

**Table 3.**
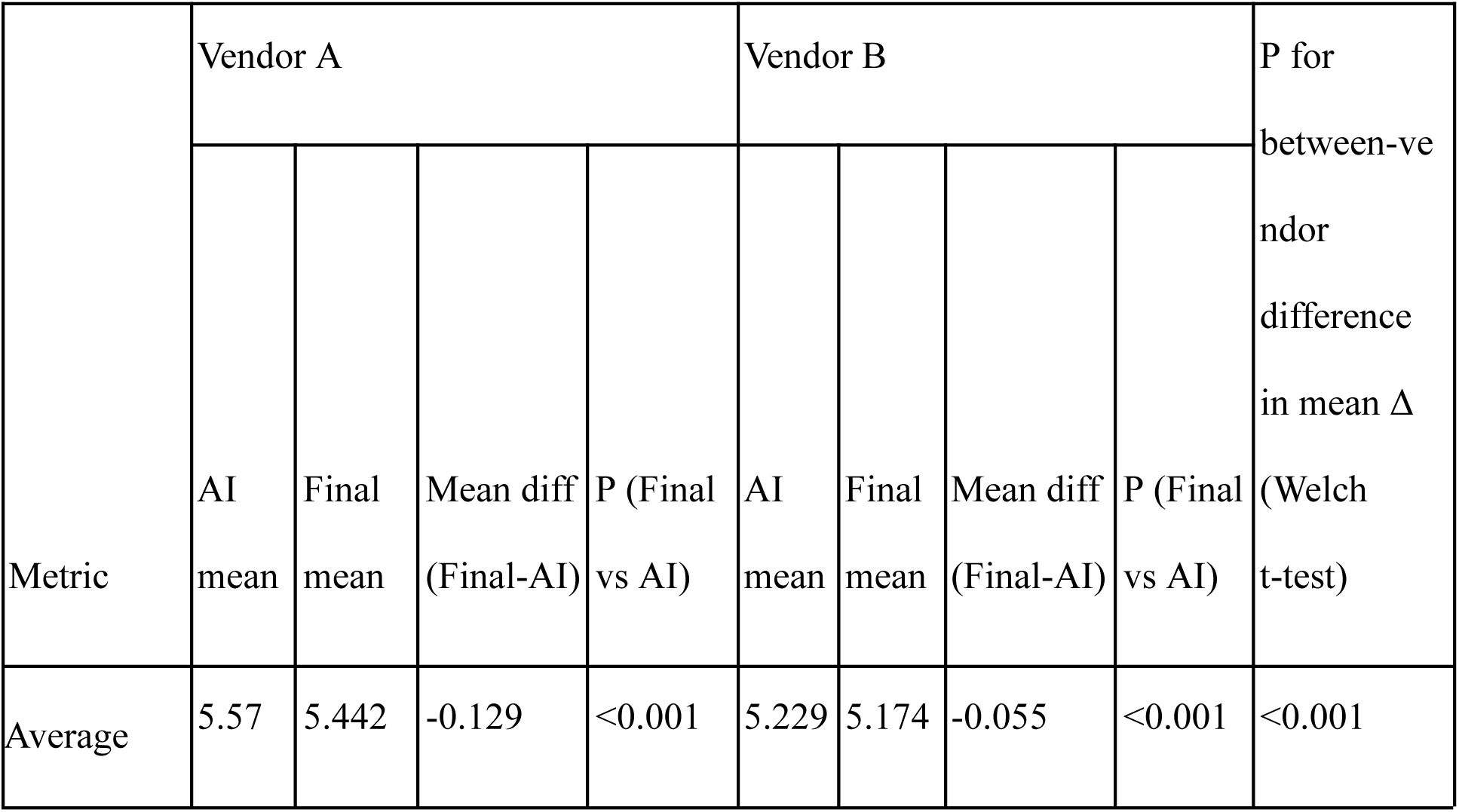
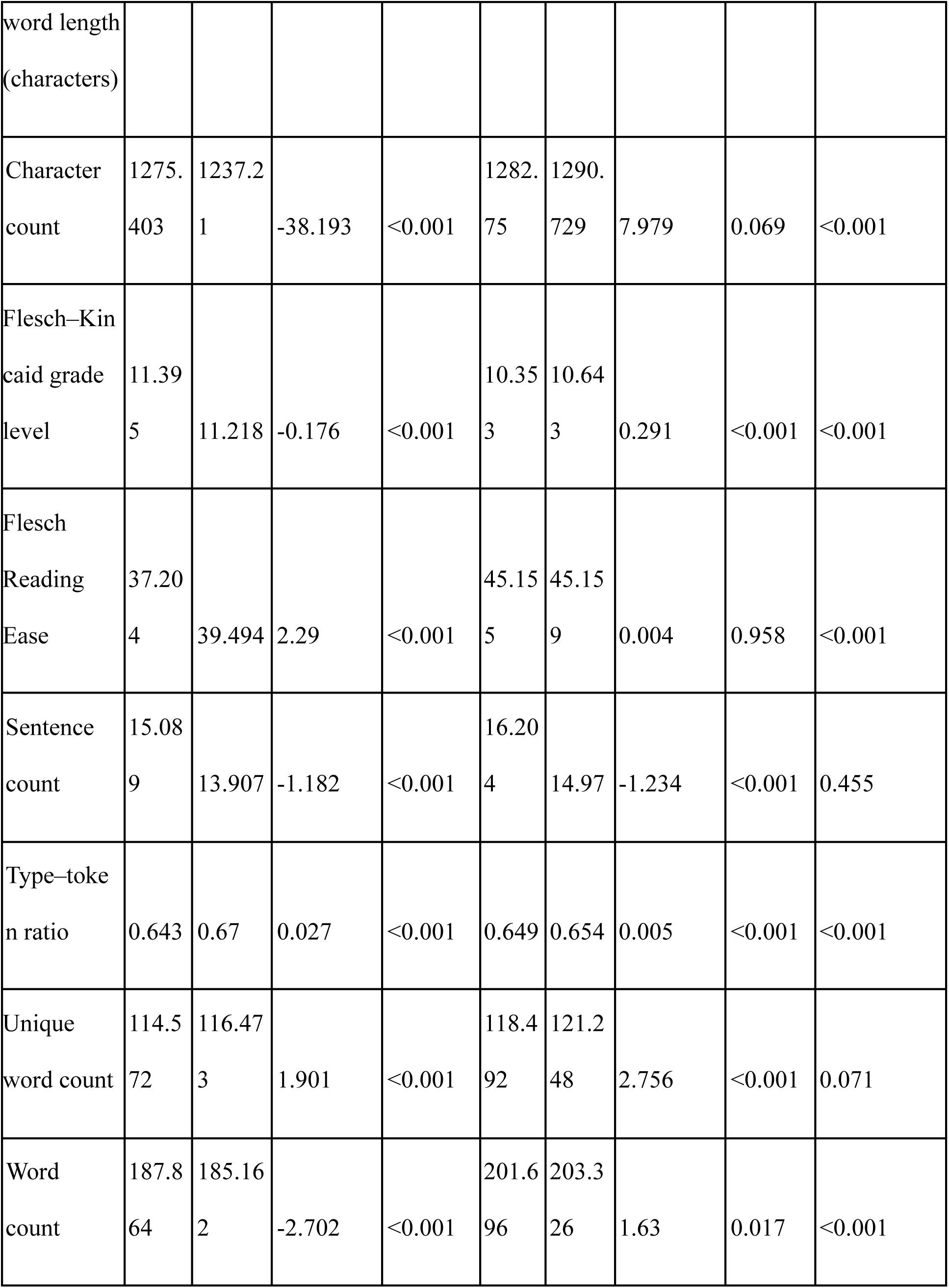
Linguistic differences comparing AI drafts and clinician-final note sections.

Across note sections, patterns of linguistic change differed by vendor (Table 4). For Vendor A, edits were most pronounced in A&P, with large decreases in character count (Δ = −164.2), word count (Δ = −17.9), and sentence count (Δ = −3.17), whereas changes in other sections were smaller and often positive (e.g., Results showed increases in character count and word count). For Vendor B, A&P changes were comparatively modest, while the largest shifts were concentrated in HPI and Results, with decreases in sentence count and increases in character count and word count. Overall, section-level differences in changes were statistically significant within both vendors (all p < 0.001).

**Table 4.**
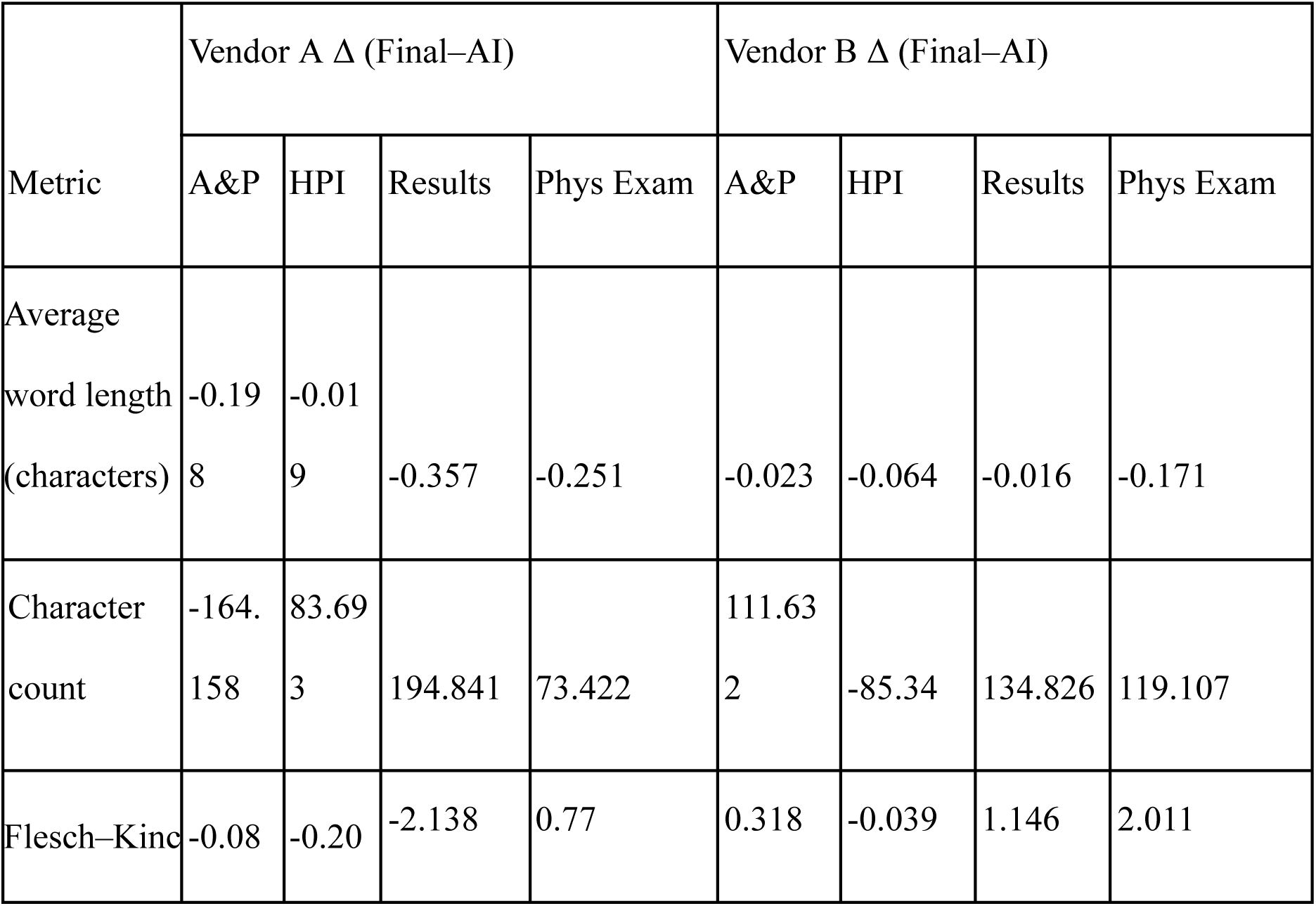
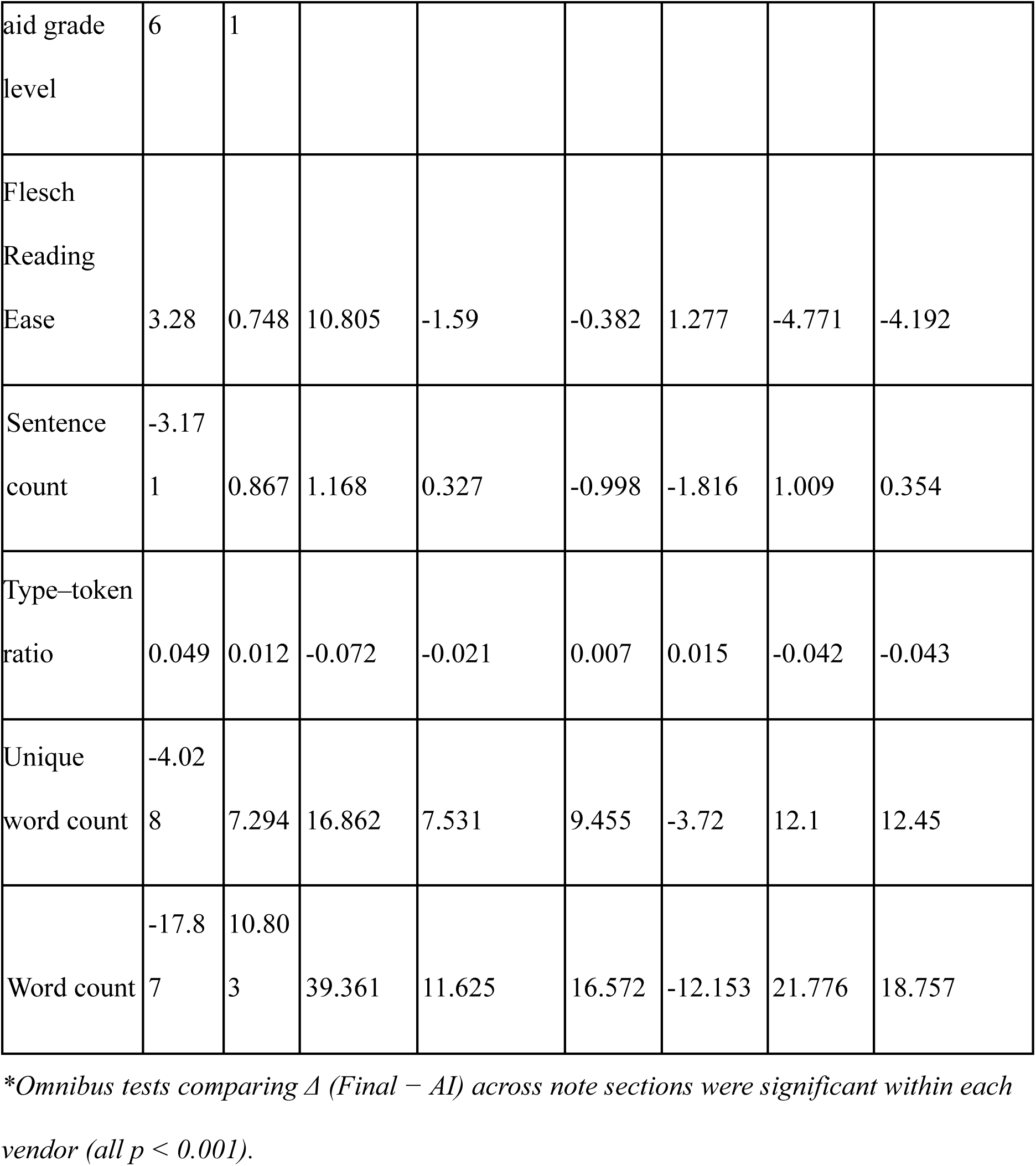
Section-Level Changes in Linguistic Metrics Between AI Drafts and Clinician-Final Note Sections.

### Which notes are more likely to have turnaround over 24 hours?

Across notes that were edited before signed (n = 20,062), 3,194 (15.9%) had turnaround over 24 hours. In vendor-adjusted models, the association between the number of AI-generated note sections within a note and long turnaround differed by vendor (Vendor × log[1+sections] likelihood-ratio test p < 0.001). For Vendor A, a higher section count was associated with greater odds of turnaround over 24 hours (log[1+sections]: β = 0.51, p = 0.010), and this association was significantly stronger for Vendor B (interaction: β = 1.06, p < 0.001). Encounter type also showed systematic differences: relative to office visits (reference), telemedicine (β = −0.52, p < 0.001) and video visits (β = −0.34, p = 0.001) had lower odds of turnaround over 24 hours; integrative medicine visits (β = 0.72, p = 0.002) and OB visits (β = 1.66, p < 0.001) had higher odds, while procedure visits were not significantly different (β = 0.12, p = 0.821). At the specialty level, family practice clinicians were less likely to have a turnaround over 24 hours (β = −3.03, p < 0.001). Several specialties had longer turnaround time, including dermatology (β = 1.46, p < 0.001) and infectious diseases (β = 2.50, p < 0.001) (Supplementary Table S3).

### Do notes signed after 24 hours have more or fewer edits?

To examine whether notes signed later had more or fewer edits, we compared total note-level edit intensity between notes signed within 24 hours versus after 24 hours. Among edited notes with valid turnaround times (n = 20,062), 16,868 notes (84.1%) were signed within 24 hours of draft generation and 3,194 (15.9%) were signed after 24 hours. Long-turnaround (>24h) was associated with lower edit loads after adjustment for encounter type, clinician group, vendor, and the log-transformed number of AI-generated note sections per note (log[1+sections]). Compared with ≤24h, >24h submissions had a reduction in log edit intensity (β = −0.269, p < 0.001). The turnaround-by-vendor interaction was not statistically significant (β = 0.073, p = 0.196). The clinician random intercept variance was estimated near zero, indicating minimal residual clustering by individual clinician behavior after these adjustments (Supplementary Table S4).

### Which sections, encounter types, and specialties show more edits?

This analysis included 33,496 edited note sections from 215 clinicians. In the adjusted model, edit intensity differed strongly by note section: compared with A&P, edit intensity was lower for HPI (β = −0.51, p = 0.002), physical exam (β = −1.36, p < 0.001), and results (β = −1.22, p < 0.001). Specialty remained associated with edit intensity relative to cardiology, with higher edit intensity in epilepsy (β = 1.60, p < 0.001), neurology (β = 1.09, p = 0.001), and physical medicine and rehabilitation (β = 0.56, p = 0.035), and lower edit intensity in family practice (β = −0.80, p = 0.003), geriatrics (β = −0.87, p = 0.019), hand surgery (β = −0.84, p = 0.004), hospitalist medicine (β = −1.60, p < 0.001), integrative medicine (β = −0.99, p = 0.002), internal medicine (β = −0.53, p = 0.027), obstetrics/gynecology (β = −0.82, p = 0.004), and ophthalmology (β = −1.18, p = 0.003). Vendor was not a significant predictor of edit intensity after adjustment (β = 0.14, p = 0.591), and encounter type was not significant in the adjusted model (all p ≥ 0.21) (Supplementary Table S5).

## Discussion

This study provides empirical evidence on ambient AI documentation usage and edit patterns among clinicians in routine ambulatory care. By linking AI drafts to clinician-final text and quantifying edits, we describe where clinicians accept AI drafts as-is, where they invest effort to revise, and how these patterns relate to turnaround time and care settings, offering a practical evaluation framework that can be reused for operational monitoring and model improvement. [31]

We found that most notes containing AI drafts (84.4%) were edited by clinicians before signing. Even though no-edit rates differed across note sections and specialties, our findings suggested substantial clinician-level heterogeneity with minimal residual clustering by specialty after accounting for section and encounter types. This suggests that the tendency to sign AI-drafted sections without edits is primarily a clinician-level phenomenon. Health systems could therefore target implementation support and workflow coaching to clinicians with unusually high or low rates of unedited submission, rather than applying training or configuration changes uniformly by specialty. [32,33] These findings also reinforce that ambient AI remains a human-in-the-loop technology, where clinician review and editing are an essential part of safe use and accountability.

When edits occurred, changes were generally modest and consistent, with the greatest editing concentrated in A&P, followed by HPI. This gradient aligns with A&P as the most information-rich note section and the primary site of clinical reasoning, risk management, and decision-making. [34] In later practice, this suggests prioritizing A&P and HPI for model refinement, such as clearer problem organization, and more specific and actionable plans with fewer generic statements, while concentrating implementation monitoring and quality checks on note sections where clinicians invest the most editing effort. [35]

Additionally, notes signed more than 24 hours after AI draft generation had lower edit intensity, suggesting that prolonged turnaround is not solely explained by extensive text revision. Longer turnaround may reflect other workflow factors (e.g., clinic scheduling, team documentation practices). Accordingly, efforts to reduce prolonged turnaround may benefit from workflow supports such as end-of-visit signing prompts and tighter integration between the ambient AI tool and EHR workflows. [36,37] In addition, because clinical notes support billing and compliance, longer AI-generated drafts should not be assumed to meet specialty-specific documentation requirements; implementation should include billing/compliance guardrails and specialty-specific guidance alongside quality monitoring. Differences in editing patterns across vendor deployments likely reflect a combination of model behavior, configuration choices, and workflow integration. Given that health systems typically implement a single ambient AI solution, our findings suggest the need for specialty- and section-specific customization and monitoring (e.g., A&P vs HPI).

This study has several limitations. Including only clinical notes from ambulatory settings within a single academic health system may limit generalizability to other institutions and clinical settings. Observed editing patterns may be shaped by local implementation factors, including system configuration, workflow integration, and clinician training and support. Because these factors vary across health systems, they may influence both the amount and type of clinician editing. Our analyses quantified the frequency and intensity of edits rather than the clinical correctness of the final notes or downstream care outcomes; therefore, we could not determine whether high no-edit use reflected high-quality drafts or missed errors, or whether higher edit levels reflected necessary clinical corrections or individual documentation style. [38]

Future work should link editing patterns to independent assessments of note quality and safety, and distinguish stylistic edits of individual clinician from clinically impactful changes to support targeted model refinement and note quality monitoring. Furthermore, prospective studies that evaluate vendor-specific performance and systematically document local implementation details could better uncover how vendor and deployment factors contribute to observed editing patterns. Overall, our proposed methods suggest an approach for health systems deploying clinical AI applications beyond ambient documentation, such as AI-drafted patient message replies: routinely track no-edit rates, edit intensity, and turnaround time to identify clinician- and content-specific friction points, and tailor implementation support to individual workflow variation rather than assuming uniform specialty-level patterns. [2,39,40]

## Conclusion

In this large-scale evaluation of two EHR-integrated ambient AI documentation systems in ambulatory settings, we found that clinicians edited the majority of AI drafted notes before signing and without edits appeared to reflect individual clinician practice style more than specialty-level norms. Documentation turnaround time was not explained by higher editing burden, as notes signed more than 24 hours after AI draft generation tended to have lower edit intensity. Section level drill-down analysis further showed that when edits occurred, revisions were generally modest and concentrated in information-dense note sections, particularly the A&P, with specialty care clinicians editing more than primary care. Vendor level differences suggest that operational monitoring and implementation support may require vendor-specific tuning. Together, these findings provide a comprehensive understanding of clinician editing behavior for ambient AI drafts to inform targeted model refinement, clinician-facing implementation support, and ongoing governance of ambient AI documentation in real-world practice.

## FUNDING STATEMENT

This study did not receive any external funding.

## COMPETING INTERESTS STATEMENT

The authors have no competing interests to declare.

## CONTRIBUTORSHIP STATEMENT

Yawen Guo (YG) led the study and contributed to the study concept and design, data acquisition, analysis, and drafting of the manuscript. Di Hu (DH) contributed to the study design and manuscript revision. Yiliang Zhou (YZ), Tianchu Lyu (TL) and Sairam Sutari (SS) provided data and administrative support and contributed to manuscript revision. Danielle Perret (DPe), Emilie Chow (EC), Steven Tam (ST), and Deepti Pandita (DPa) provided guidance through the quality improvement pilot project. Kai Zheng (KZ) provided overall supervision and guidance throughout the project. All authors critically reviewed the manuscript for important intellectual content and approved the final version.

## DATA AVAILABILITY STATEMENT

The underlying dataset contains protected health information (PHI) and is not publicly available. Summary statistics and analysis outputs are provided in the Supplementary Materials.

## Notes

### Competing Interest Statement

The authors have declared no competing interest.

### Author Declarations

Ethics committee/IRB of University of California, Irvine Institutional Review Board gave ethical approval for this work (IRB #7123).

### Summary of Updates

Vendor level data added and masked analysis done.

## References

[1] Baumann LA, Baker J, Elshaug AG. The impact of electronic health record systems on clinical documentation times: A systematic review. Health Policy 2018;122:827–36. 10.1016/j.healthpol.2018.05.014.

[2] Hassan H, Zipursky AR, Rabbani N, et al. Special Topic on Burnout: Clinical Implementation of Artificial Intelligence Scribes in Healthcare: A Systematic Review. Appl Clin Inform 2025:a-2597–2017. 10.1055/a-2597-2017.

[3] Albrecht M, Shanks D, Shah T, et al. Enhancing clinical documentation with ambient artificial intelligence: a quality improvement survey assessing clinician perspectives on work burden, burnout, and job satisfaction. JAMIA Open 2024;8:ooaf013. 10.1093/jamiaopen/ooaf013.

[4] Hundal J, Jain M, McCollom J. Ambient Artificial Intelligence in Health Care Documentation: A Review of Tools, Integration, and Clinical Implications. AI Precis Oncol 2025.

[5] Gebauer S. Benchmarking And Datasets For Ambient Clinical Documentation: A Scoping Review Of Existing Frameworks And Metrics For AI-Assisted Medical Note Generation. medRxiv 2025:2025–01.

[6] Guo Y, Wang J, Hu D, et al. Evaluating ambient artificial intelligence documentation: effects on work efficiency, documentation burden, and patient-centered care. J Am Med Inform Assoc 2025:ocaf180.

[7] Balloch J, Sridharan S, Oldham G, et al. Use of an ambient artificial intelligence tool to improve quality of clinical documentation. Future Healthc J 2024;11:100157. 10.1016/j.fhj.2024.100157.

[8] Haberle T, Cleveland C, Snow GL, et al. The impact of nuance DAX ambient listening AI documentation: a cohort study. J Am Med Inform Assoc JAMIA 2024;31:975–9. 10.1093/jamia/ocae022.

[9] Tierney AA, Gayre G, Hoberman B, et al. Ambient Artificial Intelligence Scribes to Alleviate the Burden of Clinical Documentation. NEJM Catal 2024;5. 10.1056/CAT.23.0404.

[10] Tierney AA, Gayre G, Hoberman B, et al. Ambient Artificial Intelligence Scribes: Learnings after 1 Year and over 2.5 Million Uses. NEJM Catal 2025;6. 10.1056/CAT.25.0040.

[11] Kakaday R, Herrera EZ, Coskey O, et al. The STREAMLINE Pilot – Study on Time Reduction and Efficiency in AI-Mediated Logging for Improved Note-Taking Experience. Appl Clin Inform 2025:a-2559–5791. 10.1055/a-2559-5791.

[12] Ma SP, Liang AS, Shah SJ, et al. Ambient artificial intelligence scribes: utilization and impact on documentation time. J Am Med Inform Assoc 2025;32:381–5. 10.1093/jamia/ocae304.

[13] Cao DY, Silkey JR, Decker MC, et al. Artificial intelligence-driven digital scribes in clinical documentation: Pilot study assessing the impact on dermatologist workflow and patient encounters. JAAD Int 2024;15:149–51. 10.1016/j.jdin.2024.02.009.

[14] Guo Y, Hu D, Wang J, et al. Ambient Listening in Clinical Practice: Evaluating EPIC Signal Data Before and After Implementation and Its Impact on Physician Workload. In: Househ MS, Tariq ZUA, Al-Zubaidi M, et al., editors. Stud. Health Technol. Inform., IOS Press; 2025. 10.3233/SHTI250921.

[15] van Buchem MM, Boosman H, Bauer MP, et al. The digital scribe in clinical practice: a scoping review and research agenda. NPJ Digit Med 2021;4:57. 10.1038/s41746-021-00432-5.

[16] Chishtie J, Sapiro N, Wiebe N, et al. Use of Epic Electronic Health Record System for Health Care Research: Scoping Review. J Med Internet Res 2023;25:e51003. 10.2196/51003.

[17] Pinevich Y, Clark KJ, Harrison AM, Pickering BW, Herasevich V. Interaction time with electronic health records: a systematic review. Applied clinical informatics. 2021 Aug;12(04):788–99.

[18] Nahar JK, Kachnowski S. Current and potential applications of ambient artificial intelligence. Mayo Clinic Proceedings: Digital Health. 2023 Sep 1;1(3):241–6.

[19] Cochran WG. Some Methods for Strengthening the Common χ 2 Tests. Biometrics 1954;10:417. 10.2307/3001616.

[20] Mixed Effects Logistic Regression | Stata Data Analysis Examples n.d. https://stats.oarc.ucla.edu/stata/dae/mixed-effects-logistic-regression/ (accessed December 13, 2025).

[21] Josef Perktold, Skipper Seabold, Kevin Sheppard, et al. statsmodels/statsmodels: Release 0.14.2 2024. 10.5281/ZENODO.593847.

[22] Kincaid JP, Fishburne Jr, Robert P. R, et al. Derivation of New Readability Formulas (Automated Readability Index, Fog Count and Flesch Reading Ease Formula) for Navy Enlisted Personnel: Fort Belvoir, VA: Defense Technical Information Center; 1975. 10.21236/ADA006655.

[23] Xu M, Fralick D, Zheng JZ, et al. The Differences and Similarities Between Two-Sample T-Test and Paired T-Test. Shanghai Arch Psychiatry 2017;29:184–8. 10.11919/j.issn.1002-0829.217070.

[24] Ahad NA, Yahaya SS. Sensitivity analysis of Welch’st-test. InAIP Conference proceedings 2014 Jul 10 (Vol. 1605, No. 1, pp. 888–893). American Institute of Physics.

[25] Nanda A, Mohapatra DrBB, Mahapatra APK, et al. Multiple comparison test by Tukey’s honestly significant difference (HSD): Do the confident level control type I error. Int J Stat Appl Math 2021;6:59–65. 10.22271/maths.2021.v6.i1a.636.

[26] The Myers Difference Algorithm n.d. https://www.nathaniel.ai/myers-diff/ (accessed December 13, 2025).

[27] Blizzard L, Hosmer W. Parameter Estimation and Goodness-of-Fit in Log Binomial Regression. Biom J 2006;48:5–22. 10.1002/bimj.200410165.

[28] McKnight PE, Najab J. Mann-Whitney U Test. In: Weiner IB, Craighead WE, editors. Corsini Encycl. Psychol. 1st ed., Wiley; 2010, p. 1–1. 10.1002/9780470479216.corpsy0524.

[29] Oberg AL, Mahoney DW. Linear Mixed Effects Models. In: Ambrosius WT, editor. Top. Biostat., vol. 404, Totowa, NJ: Humana Press; 2007, p. 213–34. 10.1007/978-1-59745-530-5_11.

[30] Colin Cameron A, Miller DL. A Practitioner’s Guide to Cluster-Robust Inference. J Hum Resour 2015;50:317–72. 10.3368/jhr.50.2.317.

[31] Sloss EA, Abdul S, Aboagyewah MA, et al. Toward Alleviating Clinician Documentation Burden: A Scoping Review of Burden Reduction Efforts. Appl Clin Inform 2024;15:446–55. 10.1055/s-0044-1787007.

[32] Mullankandy S, Mukherjee S, Ingole BS. Applications of AI in Electronic Health Records, Challenges, and Mitigation Strategies. 2024 Int. Conf. Comput. Appl. ICCA, Cairo, Egypt: IEEE; 2024, p. 1–7. 10.1109/ICCA62237.2024.10927863.

[33] Varghese J. Artificial Intelligence in Medicine: Chances and Challenges for Wide Clinical Adoption. Visc Med 2020;36:443–9. 10.1159/000511930.

[34] Stupp D, Barequet R, Lee I-C, et al. Structured Understanding of Assessment and Plans in Clinical Documentation 2022. 10.1101/2022.04.13.22273438.

[35] Stetson PD, Bakken S, Wrenn JO, et al. Assessing Electronic Note Quality Using the Physician Documentation Quality Instrument (PDQI-9). Appl Clin Inform 2012;3:164–74. 10.4338/aci-2011-11-ra-0070.

[36] Nguyen OT, Kunta AR, Katoju S, et al. Electronic Health Record Nudges and Health Care Quality and Outcomes in Primary Care: A Systematic Review. JAMA Netw Open 2024;7:e2432760. 10.1001/jamanetworkopen.2024.32760.

[37] Lindsay MR, Lytle K. Implementing Best Practices to Redesign Workflow and Optimize Nursing Documentation in the Electronic Health Record. Appl Clin Inform 2022;13:711–9. 10.1055/a-1868-6431.

[38] Luo Y-F, Sun W, Rumshisky A. A Hybrid Normalization Method for Medical Concepts in Clinical Narrative using Semantic Matching. AMIA Jt Summits Transl Sci Proc AMIA Jt Summits Transl Sci 2019;2019:732–40.

[39] Hu D, Guo Y, Cho HN, et al. When AI Writes Back: Ethical Considerations by Physicians on AI-Drafted Patient Message Replies 2025. 10.48550/ARXIV.2508.13217.

[40] Hu D, Guo Y, Zhou Y, et al. A systematic review of early evidence on generative AI for drafting responses to patient messages. Npj Health Syst 2025;2:27. 10.1038/s44401-025-00032-5.

