## Supplementary Table for "From Conversation to Chart: An Analysis of Clinician Edits to Ambient AI Draft Notes" (from another version of this preprint: version 1, 2026-01-06)

#### **Supplementary Table S1. Counts of AI-drafted note sections finalized without edits (“no-edit”) versus edited, by clinician specialty.**

```
=== Chi-square: Clinician_Specialty vs no_edit ===
no_edit                      False  True
Clinician_Specialty_clean
Cardiology                     470     13
Colon and Rectal Surgery        92     43
Dermatology                    141     16
Emergency Medicine              37     52
Endocrinology                  160      6
Epilepsy                       240      7
Family Practice               8859   3194
Gastroenterology               662    204
General Surgery                 22     12
Geriatric Medicine            2676   1427
Hand Surgery                  2494   1501
Head and Neck Surgery            2      1
Hematology/Oncology              5      1
Hospitalist                     80    120
Infectious Diseases             55     15
Integrative Medicine            96     37
Internal Medicine             3263   3181
Neurology                     1528     39
Neurosurgery                   292    243
Nurse Practitioner            1181    215
Ob/Gyn                         787    175
Ophthalmology                  164    285
Optometry                        7     38
Orthopaedic Surgery           3108   1370
Orthopedics                      2      2
Otolaryngology                  23    202
Physical Medicine and Rehab    540     33
Physician Assistant            906    456
Psychiatry                      14      4
Rheumatology                  4403   1510
Surgical Oncology                3      0
Urology                          2     12
Vascular Neurology              11      0
Vascular Surgery                 3      1
chi2 = 3556.942, df = 33, p = nan
```

#### **Supplementary Table S2. Mixed-effects logistic regression examining whether “no-edit” behavior is explained by individual clinician variation or specialty-level norms.**

```
                               Binomial Mixed GLM Results
==============================================================================
                                          Type Post. Mean Post. SD   SD  SD (LB) SD (UB)
------------------------------------------------------------------------------
Intercept                                    M    -1.5676   0.0124
C(Section_Name)[T.HPI]                       M     1.5108   0.0168
C(Section_Name)[T.Phys Exam]                 M     2.0126   0.0385
C(Section_Name)[T.Results]                   M     2.5875   0.0356
C(Encounter_Type)[T.Integrative Medicine]    M    -1.0936   0.1310
C(Encounter_Type)[T.OB Visit]                M    -0.6659   0.2908
C(Encounter_Type)[T.Office Visit]            M    -0.7571   0.0130
C(Encounter_Type)[T.Procedure Visit]         M    -1.1350   0.1933
C(Encounter_Type)[T.Telemedicine]            M    -0.4018   0.0697
C(Encounter_Type)[T.Video Visit]             M    -0.8153   0.0542
C(Clinician_Group)[T.Specialty Care]         M    -0.1662   0.0186
clinician                                    V     0.5872   0.0870 1.799   1.512   2.141
specialty                                    V    -3.4586   0.1665 0.031   0.023   0.044
==============================================================================
Parameter types are mean structure (M) and variance structure (V)
Variance parameters are modeled as log standard deviations

Variance components (based on log-SD parameters):
  log SD (clinician): 0.5872
  log SD (specialty): -3.4586
  SD (clinician):     1.7989
  SD (specialty):     0.0315
  Var (clinician):    3.2359
  Var (specialty):    0.0010
  Residual variance:  3.2899

Intraclass correlation coefficients (ICCs):
  ICC_clinician  = 0.496
  ICC_specialty  = 0.000
  Residual fraction = 0.504
```

Check sum ≈ 1.0: 1.000

#### **Supplementary Table S3. Editing intensity by note completion turnaround : descriptive statistics, Mann–Whitney U test, and clinician-level mixed-effects model.**

```
Edited NOTES with valid turnaround: 20188
turnaround_24h
<=24h    16955
>24h      3233

NOTE-level total edits by turnaround bucket:
turnaround_24h   count       mean        std  min  25%  50%   75%    max
         <=24h 16955.0 163.609968 210.824239  0.0 30.0 85.0 211.0 2224.0
          >24h  3233.0 139.587380 183.922829  0.0 28.0 80.0 177.0 2148.0

Mann–Whitney U test (<=24h vs >24h): U = 28792259.5, p = 5.137e-06
```

Number of clinicians with ≥100 edited NOTES: 45

```
Mixed-model NOTE-level dataset shape: (18135, 18)

Fitting MixedLM (NOTE-level) with clinician random intercept...

=== Mixed-effects model summary (NOTE-level) ===
                       Mixed Linear Model Regression Results
====================================================================================
Model:                MixedLM     Dependent Variable:     log_edit_total_tokens_note
No. Observations:     18135       Method:                 REML
No. Groups:           45          Scale:                  0.9685
Min. group size:      108         Log-Likelihood:         inf
Max. group size:      1333        Converged:              Yes
Mean group size:      403.0
------------------------------------------------------------------------------
                                          Coef.  Std.Err.   z    P>|z| [0.025 0.975]
------------------------------------------------------------------------------
Intercept                                 -0.427
C(turnaround_24h)[T.>24h]                 -0.216    0.027 -8.070 0.000 -0.269 -0.164
C(Encounter_Type)[T.Integrative Medicine]  0.006    0.583  0.011 0.992 -1.137  1.149
C(Encounter_Type)[T.OB Visit]             -0.242    0.636 -0.381 0.703 -1.490  1.005
C(Encounter_Type)[T.Office Visit]         -0.105    0.569 -0.184 0.854 -1.221  1.011
C(Encounter_Type)[T.Procedure Visit]      -0.316    0.582 -0.543 0.587 -1.456  0.824
C(Encounter_Type)[T.Telemedicine]         -0.275    0.570 -0.482 0.630 -1.393  0.843
C(Encounter_Type)[T.Video Visit]          -0.164    0.570 -0.288 0.773 -1.281  0.953
C(Clinician_Group)[T.Specialty Care]      -0.203
C(Clinician_Group)[T.Unknown]             -0.205
np.log1p(n_sections)                       1.680    0.045 37.564 0.000  1.592  1.767
Internal_Physician_ID Var                  0.000
==============================================================================


Random-effect variance from MixedLM:
  Var(clinician) (model): 0.0000
  Residual var (model):   0.9685

Clinician-level ICC (approximate, NOTE-level):
```

var_between_clin ≈ 0.846

var_within_clin ≈ 1.050

ICC_clin	≈ 0.446

Specialty-level ICC (approximate, NOTE-level):

var_between_spec ≈ 0.449

var_within_spec ≈ 1.457

ICC_spec	≈ 0.235

##### **Supplementary Table S4. Association of visit type, note section, and clinician specialty with long turnaround (>24 hours) in a binomial GLM.**

```
Total NOTES with valid turnaround: 20198
Proportion >24h: 0.16

Unique Visit_Type values: ['Office Visit' 'Telemedicine' 'Video Visit' 'Erroneous Encounter'
 'OB Visit' 'Integrative Medicine' 'Procedure Visit' 'Tumor Board']

Initial NOTE-level model sample size (N): 20198
Events (long_turnaround_24h = 1): 3233
Non-events (0): 16965

[Visit_Type] Collapsing 2 rare/perfect levels into 'Other_Visit_Type'

[Clinician_Specialty_clean] Collapsing 12 rare/perfect levels into 'Other_Clinician_Specialty_clean'

After collapsing problematic levels:
  Visit_Type: 7 levels
  Clinician_Specialty_clean: 22 levels
Model sample size (N): 20198
Events: 3233 Non-events: 16965
```

Binomial GLM for long_turnaround_24h (>24h vs ≤24h) [NOTE-level]:

```
                  Generalized Linear Model Regression Results
===============================================================================
Dep. Variable:     long_turnaround_24h   No. Observations:                20198
Model:                             GLM   Df Residuals:                    20169
Model Family:                 Binomial   Df Model:                           28
Link Function:                   Logit   Scale:                          1.0000
Method:                           IRLS   Log-Likelihood:                -7045.1
Date:                 Tue, 16 Dec 2025   Deviance:                       14090.
Time:                         17:35:27   Pearson chi2:                 2.06e+04
No. Iterations:                     20   Pseudo R-squ. (CS):             0.1664
Covariance Type:             nonrobust
==============================================================================
                                                                      coef    std err          z      P>|z|      [0.025      0.975]
------------------------------------------------------------------------------
Intercept                                                          -1.9146      0.279     -6.862      0.000      -2.462      -1.368
C(Visit_Type)[T.OB Visit]                                           1.6575      0.508      3.264      0.001       0.662       2.653
C(Visit_Type)[T.Office Visit]                                      -0.4238      0.220     -1.928      0.054      -0.854       0.007
C(Visit_Type)[T.Other_Visit_Type]                                 -21.1614   1.22e+04     -0.002      0.999    -2.4e+04     2.4e+04
C(Visit_Type)[T.Procedure Visit]                                   -0.2732      0.567     -0.482      0.630      -1.384       0.838
C(Visit_Type)[T.Telemedicine]                                      -0.9200      0.261     -3.521      0.000      -1.432      -0.408
C(Visit_Type)[T.Video Visit]                                       -0.7968      0.234     -3.410      0.001      -1.255      -0.339
C(Clinician_Specialty_clean)[T.Colon and Rectal Surgery]            1.1659      0.347      3.360      0.001       0.486       1.846
C(Clinician_Specialty_clean)[T.Dermatology]                         1.3112      0.305      4.304      0.000       0.714       1.908
C(Clinician_Specialty_clean)[T.Endocrinology]                       0.2701      0.287      0.942      0.346      -0.292       0.832
C(Clinician_Specialty_clean)[T.Family Practice]                    -1.8296      0.165    -11.100      0.000      -2.153      -1.507
C(Clinician_Specialty_clean)[T.Gastroenterology]                   -1.3422      0.258     -5.211      0.000      -1.847      -0.837
C(Clinician_Specialty_clean)[T.Geriatric Medicine]                  1.8680      0.159     11.734      0.000       1.556       2.180
C(Clinician_Specialty_clean)[T.Hand Surgery]                       -0.9037      0.177     -5.119      0.000      -1.250      -0.558
C(Clinician_Specialty_clean)[T.Infectious Diseases]                 2.6844      0.453      5.926      0.000       1.797       3.572
C(Clinician_Specialty_clean)[T.Integrative Medicine]               -2.5643      0.757     -3.387      0.001      -4.048      -1.080
C(Clinician_Specialty_clean)[T.Internal Medicine]                  -0.1476      0.160     -0.921      0.357      -0.462       0.166
C(Clinician_Specialty_clean)[T.Neurology]                          -0.3005      0.179     -1.681      0.093      -0.651       0.050
C(Clinician_Specialty_clean)[T.Neurosurgery]                       -0.1002      0.235     -0.427      0.670      -0.561       0.360
C(Clinician_Specialty_clean)[T.Nurse Practitioner]                 -1.8393      0.250     -7.359      0.000      -2.329      -1.349
C(Clinician_Specialty_clean)[T.Ob/Gyn]                             -2.7417      0.381     -7.187      0.000      -3.489      -1.994
C(Clinician_Specialty_clean)[T.Ophthalmology]                      -2.2823      0.614     -3.715      0.000      -3.486      -1.078
C(Clinician_Specialty_clean)[T.Orthopaedic Surgery]                -0.5204      0.166     -3.144      0.002      -0.845      -0.196
C(Clinician_Specialty_clean)[T.Other_Clinician_Specialty_clean]    -4.3472      1.014     -4.289      0.000      -6.334      -2.361
C(Clinician_Specialty_clean)[T.Physical Medicine and Rehab]         0.7942      0.241      3.293      0.001       0.322       1.267
C(Clinician_Specialty_clean)[T.Physician Assistant]                -0.1483      0.190     -0.781      0.435      -0.521       0.224
C(Clinician_Specialty_clean)[T.Rheumatology]                       -1.2210      0.169     -7.243      0.000      -1.551      -0.891
C(Clinician_Specialty_clean)[T.Unknown]                             0.1953      0.171      1.145      0.252      -0.139       0.529
log1p_n_sections                                                    0.9943      0.084     11.888      0.000       0.830       1.158
==============================================================================

Odds ratios (OR) for long_turnaround_24h (>24h) [NOTE-level]:
                                                                           OR  CI_lower   CI_upper
Intercept                                                        1.473941e-01  0.085303   0.254680
C(Visit_Type)[T.OB Visit]                                        5.245922e+00  1.938953  14.193073
C(Visit_Type)[T.Office Visit]                                    6.545867e-01  0.425499   1.007015
C(Visit_Type)[T.Other_Visit_Type]                                6.452343e-10  0.000000        inf
C(Visit_Type)[T.Procedure Visit]                                 7.609781e-01  0.250507   2.311664
C(Visit_Type)[T.Telemedicine]                                    3.985251e-01  0.238799   0.665087
C(Visit_Type)[T.Video Visit]                                     4.507681e-01  0.285134   0.712619
C(Clinician_Specialty_clean)[T.Colon and Rectal Surgery]         3.208880e+00  1.625478   6.334697
C(Clinician_Specialty_clean)[T.Dermatology]                      3.710621e+00  2.042433   6.741329
C(Clinician_Specialty_clean)[T.Endocrinology]                    1.310047e+00  0.746813   2.298061
C(Clinician_Specialty_clean)[T.Family Practice]                  1.604759e-01  0.116175   0.221671
C(Clinician_Specialty_clean)[T.Gastroenterology]                 2.612649e-01  0.157706   0.432826
C(Clinician_Specialty_clean)[T.Geriatric Medicine]               6.475057e+00  4.739559   8.846047
C(Clinician_Specialty_clean)[T.Hand Surgery]                     4.050741e-01  0.286601   0.572521
C(Clinician_Specialty_clean)[T.Infectious Diseases]              1.464919e+01  6.028898  35.595040
C(Clinician_Specialty_clean)[T.Integrative Medicine]             7.697132e-02  0.017452   0.339479
C(Clinician_Specialty_clean)[T.Internal Medicine]                8.627430e-01  0.630227   1.181043
C(Clinician_Specialty_clean)[T.Neurology]                        7.404472e-01  0.521622   1.051072
C(Clinician_Specialty_clean)[T.Neurosurgery]                     9.046324e-01  0.570850   1.433580
C(Clinician_Specialty_clean)[T.Nurse Practitioner]               1.589230e-01  0.097373   0.259378
C(Clinician_Specialty_clean)[T.Ob/Gyn]                           6.445898e-02  0.030520   0.136140
C(Clinician_Specialty_clean)[T.Ophthalmology]                    1.020468e-01  0.030610   0.340196
C(Clinician_Specialty_clean)[T.Orthopaedic Surgery]              5.942561e-01  0.429585   0.822050
C(Clinician_Specialty_clean)[T.Other_Clinician_Specialty_clean]  1.294312e-02  0.001776   0.094349
C(Clinician_Specialty_clean)[T.Physical Medicine and Rehab]      2.212680e+00  1.379270   3.549671
C(Clinician_Specialty_clean)[T.Physician Assistant]              8.621744e-01  0.594199   1.251003
C(Clinician_Specialty_clean)[T.Rheumatology]                     2.949435e-01  0.211960   0.410416
C(Clinician_Specialty_clean)[T.Unknown]                          1.215668e+00  0.870310   1.698072
log1p_n_sections                                                 2.702778e+00  2.294124   3.184225
```

#### **Supplementary Table S5. Predictors of heavier editing among edited AI-drafted note sections: associations by note section, encounter type, and clinician specialty.**

```
Edited notes with >0 edits: 33699
Rows after dropping missing key fields: 33699

Section_Name counts:
Section_Name
HPI          16160
A&P          14299
Phys Exam     1764
Results       1476


Visit_Type_collapsed counts:
Visit_Type_collapsed
Office Visit            29607
Video Visit              2226
Telemedicine             1058
Integrative Medicine      536
Procedure Visit           176
OB Visit                   87
Other_Visit_Type            9


Clinician_Specialty_collapsed counts:
Clinician_Specialty_collapsed
Family Practice                8818
Rheumatology                   4383
Internal Medicine              3261
Orthopaedic Surgery            3103
Geriatric Medicine             2673
Hand Surgery                   2483
Neurology                      1527
Unknown                        1460
Nurse Practitioner             1179
Physician Assistant             905
Ob/Gyn                          786
Gastroenterology                661
Physical Medicine and Rehab     540
Cardiology                      470
Neurosurgery                    292
Epilepsy                        240
Ophthalmology                   164
Endocrinology                   160
Dermatology                     141
Other_Clinician_Specialty       130
Integrative Medicine             96
Colon and Rectal Surgery         92
Hospitalist                      80
Infectious Diseases              55

Number of clinicians in model: 215


                              OLS Regression Results
==============================================================================
Dep. Variable:     log_edit_total_tokens   R-squared:                       0.265
Model:                               OLS   Adj. R-squared:                  0.265
Method:                    Least Squares   F-statistic:                     115.2
Date:                   Thu, 11 Dec 2025   Prob (F-statistic):          3.08e-114
Time:                           15:45:32   Log-Likelihood:                -53952.
No. Observations:                  33699   AIC:                         1.080e+05
Df Residuals:                      33666   BIC:                         1.082e+05
Df Model:                             32
Covariance Type:                 cluster
==============================================================================
                                                                      coef    std err          z      P>|z|      [0.025      0.975]
------------------------------------------------------------------------------
Intercept                                                           4.5032      0.367     12.264      0.000       3.784       5.223
C(Section_Name)[T.HPI]                                             -0.4955      0.162     -3.052      0.002      -0.814      -0.177
C(Section_Name)[T.Phys Exam]                                       -1.3389      0.305     -4.390      0.000      -1.937      -0.741
C(Section_Name)[T.Results]                                         -1.2276      0.217     -5.652      0.000      -1.653      -0.802
C(Visit_Type_collapsed)[T.OB Visit]                                 0.4752      0.349      1.363      0.173      -0.208       1.158
C(Visit_Type_collapsed)[T.Office Visit]                            -0.0477      0.327     -0.146      0.884      -0.689       0.593
C(Visit_Type_collapsed)[T.Other_Visit_Type]                        -0.8958      0.489     -1.833      0.067      -1.853       0.062
C(Visit_Type_collapsed)[T.Procedure Visit]                         -0.2716      0.343     -0.792      0.428      -0.944       0.401
C(Visit_Type_collapsed)[T.Telemedicine]                            -0.0668      0.345     -0.194      0.846      -0.742       0.609
C(Visit_Type_collapsed)[T.Video Visit]                              0.0601      0.315      0.191      0.849      -0.558       0.678
C(Clinician_Specialty_collapsed)[T.Colon and Rectal Surgery]       -0.5763      0.477     -1.207      0.227      -1.512       0.359
C(Clinician_Specialty_collapsed)[T.Dermatology]                     0.2692      0.171      1.576      0.115      -0.066       0.604
C(Clinician_Specialty_collapsed)[T.Endocrinology]                   0.5028      0.330      1.522      0.128      -0.145       1.150
C(Clinician_Specialty_collapsed)[T.Epilepsy]                        1.7400      0.175      9.951      0.000       1.397       2.083
C(Clinician_Specialty_collapsed)[T.Family Practice]                -0.7605      0.273     -2.788      0.005      -1.295      -0.226
C(Clinician_Specialty_collapsed)[T.Gastroenterology]               -0.6846      0.360     -1.902      0.057      -1.390       0.021
C(Clinician_Specialty_collapsed)[T.Geriatric Medicine]             -0.7356      0.257     -2.862      0.004      -1.239      -0.232
C(Clinician_Specialty_collapsed)[T.Hand Surgery]                   -0.7154      0.183     -3.900      0.000      -1.075      -0.356
C(Clinician_Specialty_collapsed)[T.Hospitalist]                    -1.6886      0.168    -10.067      0.000      -2.017      -1.360
C(Clinician_Specialty_collapsed)[T.Infectious Diseases]             0.1500      0.188      0.798      0.425      -0.219       0.519
C(Clinician_Specialty_collapsed)[T.Integrative Medicine]           -0.9862      0.315     -3.130      0.002      -1.604      -0.369
C(Clinician_Specialty_collapsed)[T.Internal Medicine]              -0.5039      0.219     -2.296      0.022      -0.934      -0.074
C(Clinician_Specialty_collapsed)[T.Neurology]                       1.2264      0.182      6.756      0.000       0.871       1.582
C(Clinician_Specialty_collapsed)[T.Neurosurgery]                   -0.9036      0.525     -1.720      0.085      -1.933       0.126
C(Clinician_Specialty_collapsed)[T.Nurse Practitioner]             -0.5223      0.430     -1.214      0.225      -1.366       0.321
C(Clinician_Specialty_collapsed)[T.Ob/Gyn]                         -0.6996      0.162     -4.306      0.000      -1.018      -0.381
C(Clinician_Specialty_collapsed)[T.Ophthalmology]                  -1.1898      0.402     -2.961      0.003      -1.977      -0.402
C(Clinician_Specialty_collapsed)[T.Orthopaedic Surgery]            -0.4867      0.202     -2.412      0.016      -0.882      -0.091
C(Clinician_Specialty_collapsed)[T.Other_Clinician_Specialty]      -0.6160      0.410     -1.503      0.133      -1.419       0.187
C(Clinician_Specialty_collapsed)[T.Physical Medicine and Rehab]     0.5659      0.264      2.146      0.032       0.049       1.083
C(Clinician_Specialty_collapsed)[T.Physician Assistant]            -0.7141      0.539     -1.324      0.185      -1.771       0.343
C(Clinician_Specialty_collapsed)[T.Rheumatology]                    0.4809      0.474      1.014      0.310      -0.448       1.410
C(Clinician_Specialty_collapsed)[T.Unknown]                         0.0546      0.255      0.214      0.830      -0.445       0.555
==============================================================================
Omnibus:                      716.966   Durbin-Watson:                   1.718
Prob(Omnibus):                  0.000   Jarque-Bera (JB):              745.970
Skew:                          -0.353   Prob(JB):                    1.03e-162
Kurtosis:                       2.815   Cond. No.                         94.1
==============================================================================
```
